# Perceptions of the implementation of a case–creation methodology in a health management course among medical students: a mixed–methods study

**DOI:** 10.1101/2025.10.25.25329949

**Authors:** Leonardo Rojas–Mezarina, Stefan Escobar–Agreda, Javier Silva–Valencia, Juan J. Quispe-Gamarra, Héctor Espinoza–Herrera, Katherine A. Pérez–Acuña Medina, Ada Pastor Goyzueta

## Abstract

**Background:** In Peru, the Universidad Nacional Mayor de San Marcos involved teachers and students in the creation of management cases in the Rural Urban Marginal Health Service (SERUMS). However, their perception of the implementation of this methodology is not known.

**Objective:** To evaluate the perception of students and teachers on the implementation of a case-creation methodology in the Health Management course at UNMSM.

**Methods:** Mixed methods design. Questionnaires (quantitative phase) and focus groups (qualitative phase) were used to explore the perceptions of students and teachers on the implementation of a case-creation methodology in the Health Management course at UNMSM during 2019.

**Results:** Of the 182 surveyed students, 61% found the created cases interesting and 63% stated that the methodology encourages the use of critical thinking. Qualitatively, the possibility to learn about real SERUMS cases and work collaboratively to find innovative solutions was highlighted. However, several mentioned that this led to an academic overload and emphasized a lack of information on the topic. Teachers mentioned that the methodology promoted learning in management, though they recognized challenges in its implementation due to the limited experience and training of teachers and students.

**Conclusions:** The case-creation methodology is perceived as an innovative and beneficial and educational strategy for teaching managerial aspects in SERUMS. However, an adequate process of training, planning, and organization is suggested for its effective implementation.

## Introduction

Case-Based Learning (CBL) is a pedagogical strategy that uses the narration of a situation or simulated case containing problems and challenges within a specific context, where participants must find solutions based on well-founded decisions applying theories specific to a field of specialization (1,2). This strategy has been adopted in numerous educational institutions, showing significant improvements in students’ critical thinking, research skills, and decision-making (3– 5). Moreover, CBL has been shown to facilitate the connection between theoretical and practical knowledge, foster motivation, improve exam preparation, and stimulate critical reflection (6–8).

In Peru, a notable deficit in medical education is the acquisition of knowledge and skills in Health Management, which are crucial for professionals participating in the Rural and Marginal Urban Health Service (SERUMS). Indeed, although a large proportion of physicians performing this service carry out administrative duties during this period, there is a general perception of insufficient preparation in management competencies among recent medical graduates (9,10). While CBL could help mitigate this educational gap, the lack of model cases limits the teaching of management aspects within SERUMS, since in Peru, the application of this educational methodology has been mainly restricted to clinical practice (11,12) and hospital management (13).

The creation of cases for CBL involves a meticulous cognitive and organizational process, in which cases must have appropriate coherence and narrative structure and must be designed based on specific teaching objectives (14,15). Given these particularities, the Health Management course for medical students at the Universidad Nacional Mayor de San Marcos (UNMSM) decided to involve both specialized faculty members and medical students in the creation of cases related to management aspects of the SERUMS program. This initiative aimed not only to promote the future application of CBL in this area but also to establish a formative methodology that fosters collaborative participation between faculty and students. Despite its potential, the perception of how this methodology is implemented in practice remains unknown. This study aims to evaluate students’ and faculty members’ perceptions of the implementation of a case-creation methodology in the Health Management course at UNMSM.

## Methods

### Study design and population

A sequential explanatory mixed-methods design was followed, according to Creswell (16), in which quantitative information was first collected through virtual surveys, followed by qualitative data collection through focus groups.

### Population and sample

For the quantitative phase, participants included sixth-year medical students who took part in the case-creation methodology as part of the Health Management course at the Faculty of Medicine of the Universidad Nacional Mayor de San Marcos during 2019. All students enrolled in the course were invited to participate in the study. For the qualitative phase, both students and faculty members who participated in this course were included.

### Case-creation methodology

The design of the case-creation methodology involved a series of sessions with the instructors of the Health Management course, composed of physicians specialized in Health Management and Administration. During these sessions, the definition of a case according to the CBL strategy and the evaluation criteria for the created cases were explained, and the most suitable approach for implementing this methodology was decided. This included the formation of working groups composed of students under the guidance and supervision of a course instructor.

At the beginning of the course, students received a workshop explaining the CBL strategy and the step-by-step process for case creation. All this information was also shared through detailed guides that included examples of model cases developed for other medical specialties. Throughout the course, ten group sessions were coordinated between instructors and students, during which they had to create the case and its corresponding solution.

The case-creation process consisted of defining the setting and specific health management issue to be addressed, incorporating a character involved in this issue, describing the data to be used for case resolution, and formulating questions at the end of the case related to the learning objectives based on the CBL strategy. At the end of the course, each group presented and explained the cases they had developed with their instructors, describing their characteristics, narratives, and proposed solutions.

### Variables and instruments

In the quantitative phase, students’ perceptions of their participation in the intervention were assessed using an adaptation of the questionnaire designed by Dalal and Kaja (17), originally developed to evaluate the CBL strategy. The questionnaire assessed various aspects of the case-creation methodology, including planning, relevance, learning support, interaction, and related dimensions. Responses were categorized on a five-point scale (1 = strongly disagree, 2 = disagree, 3 = neither agree nor disagree, 4 = agree, 5 = strongly agree) based on participants’ perceptions.

For the qualitative phase, focus groups were conducted with course instructors to explore their perceptions regarding the implementation of the case-creation methodology. In the case of students, an open-ended question was added at the end of the survey, asking why they would or would not recommend the incorporation of the case-creation methodology into the Health Management course in future semesters.

### Analysis plan

For the quantitative phase, a descriptive analysis was conducted. Numerical variables were summarized using means and standard deviations, while categorical variables were summarized using frequencies and proportions. Analyses were performed using Stata version 17.

For the qualitative phase, a qualitative grounded theory analysis was performed. Analyses were conducted using the software, Atlas.ti.

### Ethical considerations

The present study was approved by the Research Ethics Committee of the Faculty of Medicine of the Universidad Nacional Mayor de San Marcos. Its development adhered to the principles of the Helsinki and Taipei Declarations.

## Results

### Quantitative results

Of the 214 students enrolled in the Health Management course in 2019, 182 (85.5%) agreed to participate in the study and were ultimately evaluated. Regarding their perception of the case-creation methodology, 61% considered the cases created to be interesting, 63% mentioned that the case-creation process allowed them to use their critical thinking skills, 53% stated that creating cases would help them in future applications of this knowledge, and 50% indicated that it promoted interaction among students. Additionally, 71% considered that the instructors’ role was important for the collaborative creation of cases.

On the other hand, only 29% of students reported that the cases were well-organized and well-planned. Fewer than half stated that case creation improved their independent learning and communication skills (39%) or that they would recommend the use of case creation for future cohorts of the course (44%). Moreover, 47% did not believe that case creation improved their exam performance, and the majority (77%) considered that case creation increased their workload.

**Figure 1.**
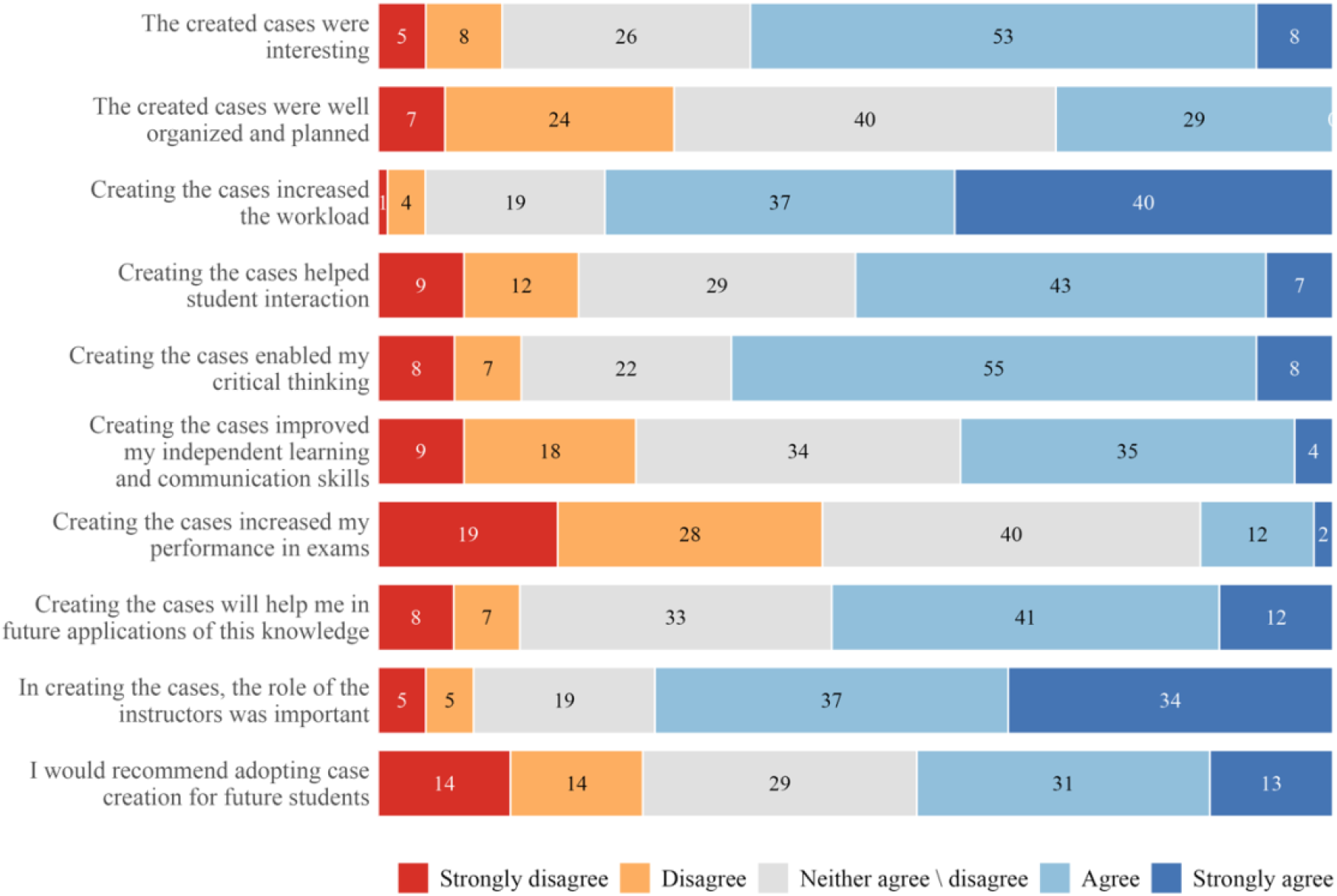
Distribution of student responses to the perception survey on the case-creation methodology.

### Qualitative results

Regarding the open-ended comments provided by students about this methodology, they highlighted that it offered them the opportunity to learn about real situations experienced by other physicians in managerial roles during the Rural and Marginal Urban Health Service (SERUMS), within the context of the Peruvian health system. This experience allowed them to become aware of the working conditions they would face in the future, especially because they were nearing the completion of their studies.

> *“It helped us become aware of the problems that arise in SERUMS and other healthcare centers, as well as to understand the reality and conditions of the health system*.*”*

Students also viewed this experience as an opportunity to apply critical thinking, put into practice the knowledge acquired throughout their university years, and gain new insights related to management. They also saw it as a way to promote collaborative work among peers to find solutions to future problems that could hinder their work as physicians in rural or underserved urban areas.

> *“We were able to discuss, as a group, the problematic situations we might face during SERUMS, and that helps us be better informed about the correct way to act to avoid problems*.*”*

On the other hand, students identified difficulties in participating in this methodology, such as their limited understanding of administrative or managerial issues that could arise in primary healthcare centers in rural or underserved urban areas of the country. They noted the lack of bibliographic sources and of experienced alumni performing their SERUMS who could share real-life insights.

> *“We’re not fully aware of the real problems in primary care centers. As students, we haven’t had much contact with health posts, etc. (…)”*

Additionally, some students indicated that the course represented an academic overload, as they were simultaneously preparing for the internship selection exam at the Social Security system (EsSalud). Others were not particularly interested in the course, which delayed group progress and depended heavily on their instructor’s evaluation criteria.

> *“The biggest issue is that, as you know, in the sixth year, most students are focused on preparing for the internship or the EsSalud exam, so they’re not really interested in this type of course*.*”*

Students also mentioned that the theoretical training provided was insufficient to fully understand the structure and objectives of the cases and that some instructors lacked in-depth knowledge about the methodology.

Among the students’ recommendations, they suggested better planning of activities involving student participation from the first day of class. They emphasized that both students and instructors should receive the same training to ensure a shared understanding of key concepts.

> *“I think there should be a clearer explanation of what exactly is expected as a final product, because each instructor seems to have their own criteria*.*”*

They also suggested providing examples of previous cases created by other students or graduates. If possible, they proposed inviting a physician with management experience to share real-world problems and solutions, including their positive or negative consequences.

> *“It would be more helpful to have real stories from current or former SERUMS participants and discuss them as cases. (…) I would even suggest recording their testimonies and showing them as videos in class*.*”*

### Faculty perceptions

Instructors shared their perspectives on the implementation of the case-creation methodology through a focus group. Among the positive aspects, they highlighted the analytical process students undertook when reflecting on others’ managerial experiences, which fostered discussion and encouraged original problem-solving. They also noted that while students were already familiar with clinical cases, managerial cases were new and therefore promoted critical analysis, research, integration, and synthesis to solve the cases.

> *“I found it interesting because, as physicians, we’ve always worked with clinical cases and case presentations. This was innovative, applying it to the management and administrative side, especially cases related to SERUMS*.*”*

They also found it novel how the course content evolved, since many students had not perceived Health Management as an important subject in their medical education. However, after engaging with real cases from previous SERUMS experiences, greater student interest was observed. Furthermore, instructors noted that this methodology helped them instill in students a better understanding of the healthcare reality they would soon face, while also providing them with tools to address similar issues.

> *“When we learned about this (case-creation methodology), we found it very interesting — to have cases from the primary care level, which are not easy to come by*.*”*

Regarding negative aspects, instructors acknowledged that creating cases was challenging, as they were not familiar with developing such materials. Previously, they used pre-existing cases for class discussions, but now they had to create new ones with students and provide evidence-based solutions.

> *“These were managerial cases, but before, we were given pre-made cases to analyze with students. It’s different when you have to create the case yourself from scratch*.*”*

Another difficulty mentioned was the students’ lack of experience with management-related problems, which forced them to rely on limited bibliographic sources. Additionally, addressing the case problems often required consulting Peruvian health regulations.

> *“The main challenge was having students put themselves in a situation they hadn’t yet experienced. There’s also a lack of published literature on first-level management cases with sufficient depth and analysis to support the course*.*”*

*They also noted that the time allocated for training in this methodology was insufficient to establish clear and precise objectives, which in turn affected their ability to respond effectively to students’ questions*.

> *“I need to learn more about case creation, because the two sessions we had went by too quickly*.*”*

Finally, instructors emphasized the importance of encouraging students to communicate with physicians currently performing SERUMS, so that their experiences could provide a broader perspective on real-world managerial challenges in rural or underserved settings. They also proposed publishing the cases created in an open repository to allow future students access to these materials.

> *“They should connect with current SERUMS participants so they can gather real data and use it throughout the course*.*”*

## Discussion

The implementation of the case-creation methodology focused on managerial aspects within the SERUMS program was perceived as a novel yet challenging educational experience by both students and instructors of the Health Management course at UNMSM. It was noted that this methodology offers several features that enhance and motivate student learning, although it also presents certain limitations in its implementation which, if not properly addressed, may restrict its potential as an educational tool.

Among the positive aspects, students highlighted the opportunity to learn about real-life situations in rural and underserved urban areas that involve administrative or managerial challenges— mainly at the primary healthcare level within the current context of the Peruvian health system. This promoted awareness among students about the need to develop competencies in health management, which could help reduce the training gap in managerial skills perceived among newly graduated physicians in Peru (9), particularly in preparation for their upcoming work in rural or underserved settings as part of the SERUMS program.

Another positive aspect was the encouragement of collaborative work and the application of critical thinking among students in order to find innovative, systematic, and well-reasoned solutions to new problems. These characteristics, which are also attributed to the Case-Based Learning (CBL) strategy (5), suggest that the case-creation methodology may represent a distinct teaching approach that fosters teamwork, an essential component in the field of health management (18).

Regarding the negative aspects, the lack of student knowledge about managerial situations occurring at the primary healthcare level was identified as a major limitation for developing realistic and applicable cases. The limited understanding of health management is linked to the deficiencies in undergraduate medical training, which is heavily focused on solving clinical problems in hospital settings and places little emphasis on managerial issues at the primary care level, ultimately affecting performance during the SERUMS program (19). Furthermore, the available literature is mostly restricted to personal experiences and recommendations from physicians who have completed their SERUMS (20), further constraining the resources available for developing relevant and useful cases.

Finally, difficulties were noted in the organization and training of both students and instructors for the proper implementation of the methodology. Multiple group sessions were required for students, guided by instructors, to develop cases aligned with the CBL structure, including the definition of the problem, the resolution process, and learning objectives. Consequently, many students perceived this activity as an additional academic burden on top of their existing workload.

## Conclusions

The case-creation methodology is perceived as a promising and beneficial educational strategy that promotes learning about managerial aspects in primary healthcare, while fostering collaboration and critical thinking among medical students. However, its effective implementation faces challenges such as students’ limited preparation for primary care settings, insufficient bibliographic resources, and its perception as an added academic workload. To maximize the effectiveness of this methodology as a pedagogical tool, it is crucial to strengthen instructor training, improve planning and organization, and ensure the availability of adequate educational resources, thereby enhancing the learning outcomes of participating students.

## Data Availability

All data produced in the present study are available upon reasonable request to the authors

## Notes

**Conflict of interest:** The authors declare no conflicts of interest.

### Competing Interest Statement

The authors have declared no competing interest.

### Funding Statement

This study did not receive any funding

### Author Declarations

The present study was approved by the Research Ethics Committee of the Faculty of Medicine of the Universidad Nacional Mayor de San Marcos.

